# Case Fatality of Leptospirosis in the Dominican Republic, 2012–2026: A 14-Year National Surveillance Analysis

**DOI:** 10.64898/2026.09.01.26361950

**Authors:** José Javier Sánchez, Lisette V. Alcántara, David De Luna, O. Alejandro Aleuy, McKenzie Bybee Bellon, José Luis Cruz Raposo, Timothy De Ver Dye

## Abstract

**Background:** Case fatality reflects the quality and timeliness of clinical care for leptospirosis, yet no study has examined it at a national level in the Dominican Republic (DR), where leptospirosis is endemic. We describe the case fatality rate (CFR) of leptospirosis in the DR between 2012 and 2026 and identify associations with mortality.

**Methods:** We conducted an analytical cross-sectional study, using national surveillance records merged with a discharge-condition extract via a composite key. We calculated CFR with Wilson 95% confidence intervals among 5,412 valid cases (suspected, probable, or confirmed) reported from January 2012 through June 2026. We compared proportions with Pearson’s chi-square test, assessed annual trend with ordinary least squares linear regression, and fitted multivariable logistic regression models to account for confounding.

**Results:** Overall CFR was 8.5% (460/5,412), with no significant annual trend (p=0.372). CFR was higher in men than women (p < 0.001) and increased significantly with age (p < 0.001). Male gender (OR: 1.79; 95%CI: 1.18–2.73) and pre-existing comorbidity (OR: 1.76; 95% CI: 1.21– 2.57) were independent predictors of death. Clinical complications were the strongest predictor in the adjusted model (OR: 3.16; 95%CI: 2.17–4.61), attenuating the gender effect. CFR varied widely by province (2.43–22.22%) and correlated negatively with incidence at the province level (p = 0.066).

**Conclusions:** Leptospirosis case fatality is concentrated among men, people with comorbidity, and those who develop clinical complications. These national, long-term findings can help prioritize clinical and surveillance resources as extreme weather events are expected to intensify across the Caribbean.

## Introduction

Leptospirosis is a bacterial zoonosis caused by pathogenic spirochetes of the genus Leptospira, transmitted mainly through contact with water or soil contaminated by the urine of reservoir animals (1–3). Clinical presentation ranges from a self-limited febrile illness to severe forms such as Weil’s syndrome (jaundice, acute renal failure, hemorrhagic diathesis) and severe pulmonary hemorrhage syndrome, the latter with case fatality exceeding 50% among those who develop it (4–5).

The WHO Leptospirosis Burden Epidemiology Reference Group estimated approximately 1.03 million severe cases and 58,900 deaths annually worldwide, with nearly 80% of the estimated 2.9 million annual DALYs occurring in men (1,6). The greatest burden falls on tropical low- and middle-income regions, where surveillance limitations likely lead to systematic underestimation (1,6–7).

The reference standard for serological confirmation, the microscopic agglutination test (MAT), requires trained personnel, a locally representative live-strain panel, and paired sera, making it impractical for timely clinical diagnosis in most endemic settings; a recent systematic review and meta-analysis confirmed substantial heterogeneity in MAT sensitivity across settings (8). Studies comparing passive surveillance with active case-finding have found that routine diagnostic pathways capture only 50-80% of actual hospitalized cases depending on whether diagnosis relies on clinical criteria or MAT confirmation (9), a limitation directly relevant to the suspected/probable/confirmed classification used in this study.

Latin America and the Caribbean account for a disproportionate share of the global leptospirosis burden, with approximately one-third of globally reported outbreaks between 1970 and 2012 occurring in the region (10–11) and DALY-based estimates ranking it among the highest-risk regions worldwide (1,6). A scoping review of insular Caribbean literature published between 2000 and 2022 identified only 16 studies with usable data from 15 of 27 countries and territories, concluding that substantial gaps remain in surveillance and in understanding of environmental drivers, even as urbanization and extreme weather increasingly shift transmission from a predominantly rural to an urban pattern (12).

The paradigm of urban leptospirosis in the region was established by prospective cohort studies in Salvador, Brazil, which quantified an annual infection risk exceeding 3% among slum residents and identified household elevation, drainage, and socioeconomic status as independent risk determinants (13–18). A similar pattern has been documented in the Dominican Republic: a 2021 national seroepidemiology survey confirmed considerable population-level seroprevalence (19), and a geospatial analysis of two provinces found that the Ministry of Public Health’s surveillance system received 2,860 case notifications between 2013 and 2023, concentrated in areas of lower population density, lower gross domestic product, and greater distance from health facilities, characteristics typical of structural poverty (11).

Outside Latin America and the Caribbean, countries sharing a tropical climate, middle-income economy, and surveillance infrastructure of comparable structure offer a useful reference framework. In Thailand, a prospective cohort study in Khon Kaen identified pulmonary hemorrhage, complicated acute renal failure, and multiple organ failure as the leading causes of death among hospitalized patients, with a mortality rate of 1.4 per 100 patient-days of observation (20).

In Sri Lanka, a systematic review adjusting for underdiagnosis and underreporting estimated an annual hospitalized incidence of 52.1 cases per 100,000 population and approximately 730 deaths per year, several times higher than unadjusted surveillance figures (9); a study optimizing the MAT strain panel found positivity of only 11% among over a thousand clinically suspected patients, illustrating the diagnostic challenge even with dedicated reference infrastructure (21). Colombia, with a mandatory laboratory surveillance system structurally similar to the Dominican one, offers the most direct point of comparison: an analysis of 201 laboratory-confirmed cases between 2015 and 2020 found a case fatality of 8.5%, a male proportion of 85.6%, and 43.3% of cases classified as severe, with renal failure, liver failure, and multiple organ failure as the most frequent complications (22), a profile remarkably convergent with the Dominican cohort analyzed here.

Despite this growing body of evidence on incidence, environmental determinants, and risk factors for leptospirosis in the Dominican Republic and comparable settings, to our knowledge no analysis of the disease’s case fatality rate (CFR) in the country has covered an extended period and the entire national territory. This omission is relevant because case fatality, unlike incidence, speaks directly to the quality of clinical care, diagnostic and therapeutic timeliness, and the identification of population subgroups at greater risk of a fatal outcome. The present study aims to describe the case fatality of leptospirosis reported to the Dominican Republic’s national epidemiological surveillance system between 2012 and 2026, and to determine the clinical, demographic, and geographic factors associated with a fatal outcome.

## Materials and Methods

### Study Design and Setting

We conducted a cross-sectional observational epidemiological study, with a temporal trend component, aimed at characterizing the case fatality rate (CFR) of leptospirosis and identifying the clinical, demographic, and geographic factors associated with a fatal outcome. We used records from the national epidemiological surveillance system of the Dominican Republic, for the period from January 2012 through June 2026 (14 complete calendar years plus a partial final half-year). Descriptive statistics and stratified case fatality estimates use the full period; the annual trend regression (below) is restricted to the 14 complete calendar years (2012–2025) to avoid biasing the estimated slope with a partial final year, and this restriction is noted again where the trend analysis is described. We disaggregated the analysis by province (22) and by health region of the National Health Service (SNS: I Valdesia, II Cibao Norte, III Cibao Nordeste, IV Enriquillo, V Este, VI El Valle, VII Cibao Occidental, VIII Cibao Central, and O Metropolitana).

### Data Source and Case Definition

Case data came from the Epidemiological Surveillance Information System (SIP-0276FA51, Leptospirosis, 2010–2026) of the Ministry of Public Health (MSP), through the General Directorate of Epidemiology (DIGEPI) (23): 5,412 records classified as suspected, probable, or confirmed, after excluding discarded records (n=186) and those pending classification or missing this variable (n=2,827).

We incorporated into this cohort the clinical outcome variable “condicion de egreso” (discharge condition: alive/deceased), provided by the MSP in an additional extract of the surveillance database (SIP-0276FA51_CONDICION_DE_EGRESO). Because neither database had a unique case identifier, we constructed the merge using a composite key built from 53 variables shared between both extracts (date of symptom onset, notification date, gender, age, province, final laboratory result, among others). This procedure allowed us to assign discharge condition to all 8,425 records in the original working database, including a second reconciliation pass for 10 records in which missing province and region-of-residence data in the discharge-condition extract prevented a match on the first pass. Of the 5,412 valid cases, 460 (8.5%) had death as their discharge condition.

### Population Denominators

We did not require population denominators for the case fatality analyses, since CFR is defined as a proportion calculated directly over total cases (numerator: deaths; denominator: valid cases), not as a population rate. National Statistics Office (ONE) denominators were used only in a complementary capacity, to contrast the geographic pattern of case fatality with that of incidence by province (23).

### Study Variables

The primary outcome variable was discharge condition, dichotomized as alive/deceased. As independent variables we included: gender, age group (nine categories), province and health region of residence, final case classification, presence of pre-existing comorbidities, presence and number of clinical complications documented during the course of illness, and the intervals between symptom onset and medical care, and between symptom onset and notification to the surveillance system, the latter two used as indicators of care timeliness.

### Statistical Analysis

#### Calculation of Case Fatality

We calculated overall and subgroup-specific CFR (by year, gender, age group, final classification, province, health region, comorbidity, and complications) as the ratio of deaths to valid cases in the corresponding subgroup, expressed as a percentage. We estimated the 95% confidence interval for each CFR using the Wilson method for binomial proportions, more appropriate than the classic normal-approximation method when some categories have proportions near 0 or 100% and small sample sizes, a situation present in several provinces with comparatively few cases.

#### Comparison of Proportions and Medians

We assessed differences in case fatality between categories using Pearson’s chi-square test. We calculated the male-to-female case fatality risk ratio as the ratio of the gender-specific proportions. We compared care-timeliness intervals (median days) between those who died and survivors using the Mann-Whitney U test, given the skewed distribution of these variables.

We assessed the trend in annual case fatality (2012–2025, complete years) using ordinary least squares linear regression of the annual percentage CFR on calendar year.

#### Multivariable Logistic Regression Models

We fitted two multivariable logistic regression models to identify independent predictors of mortality. Model A included gender, age group (reference: 20–29 years), comorbidity, and year of occurrence (centered on the median year) as covariates. Model B additionally incorporated the presence of clinical complications, to explore whether the excess mortality observed by gender and comorbidity was mediated by the occurrence of complications. In both models we grouped age categories under 10 years into a single category (0–9 years), since modeled separately they produced quasi-perfect separation (absence of deaths) that prevented estimation of finite coefficients. The clinical severity field recorded in the surveillance system showed a null association with discharge condition (no case with a recorded severity value had a fatal outcome), suggesting this variable is preferentially documented for patients who survive to hospital discharge; for this reason we did not use it as a severity covariate in the multivariable models.

We estimated coefficients using the iteratively reweighted least squares algorithm (IRLS, Newton– Raphson), with standard errors derived from the Fisher information matrix, and expressed results as odds ratios with 95% confidence intervals and Wald test p-values.

All statistical tests were two-sided, with statistical significance defined as p < 0.05. Data management, descriptive analyses, and statistical analyses were performed using IBM SPSS Statistics (version 29).

### Ethical Considerations

The protocol for this study (“Leptospirosis in the Dominican Republic, 2000–2026: National Surveillance Trends,” protocol IRB2606182) was reviewed by the Florida Atlantic University Institutional Review Board, which determined it exempt from federal regulation under 45 CFR 46.104(2)(ii) on June 22, 2026, given its nature as a secondary analysis of already collected, de-identified surveillance data. This is the same protocol and exemption determination covering the companion incidence manuscript, since both articles derive from the same cohort and the same anonymized dataset provided by the MSP, with no access to names, identification documents, or any other information that could directly identify patients.

## Results

### Cohort Characteristics and Overall Case Fatality

Of the 5,412 valid leptospirosis cases identified between 2012 and June 2026, 460 (8.5%) had death as their discharge condition (Table 1). The proportion of deaths that were male (82.4%) exceeded the proportion of male non-fatal cases (71.8%), consistent with the higher case fatality in men described below (male:female risk ratio = 1.75). Comorbidity and clinical complications were also more frequent among those who died: 11.3% of deaths had at least one recorded comorbidity, versus 6.2% of survivors, and 25.9% of deaths had at least one documented clinical complication, versus 7.4% of survivors.

**Table 1.** Cohort characteristics, by vital status (alive/deceased), N = 5,412.

| Characteristic | Alive | Deceased | Total |
| --- | --- | --- | --- |
| N | 4952 | 460 | 5412 |
| Gender |  |  |  |
| Male | 3558 (71.8%) | 379 (82.4%) | 3937 (72.7%) |
| Female | 1394 (28.2%) | 81 (17.6%) | 1475 (27.3%) |
| Age group |  |  |  |
| <1 year | 24 (0.5%) | 1 (0.2%) | 25 (0.5%) |
| 1 to 4 years | 168 (3.4%) | 1 (0.2%) | 169 (3.1%) |
| 5 to 9 years | 300 (6.1%) | 6 (1.3%) | 306 (5.7%) |
| 10 to 19 years | 1209 (24.4%) | 90 (19.6%) | 1299 (24.0%) |
| 20 to 29 years | 1094 (22.1%) | 132 (28.7%) | 1226 (22.7%) |
| 30 to 39 years | 646 (13.0%) | 72 (15.7%) | 718 (13.3%) |
| 40 to 49 years | 524 (10.6%) | 42 (9.1%) | 566 (10.5%) |
| 50 to 59 years | 453 (9.1%) | 57 (12.4%) | 510 (9.4%) |
| >60 years | 534 (10.8%) | 59 (12.8%) | 593 (11.0%) |
| Final classification |  |  |  |
| Confirmed | 476 (9.6%) | 47 (10.2%) | 523 (9.7%) |
| Probable | 1284 (25.9%) | 164 (35.7%) | 1448 (26.8%) |
| Suspected | 3192 (64.5%) | 249 (54.1%) | 3441 (63.6%) |
| Comorbidity (available data) |  |  |  |
| Yes, >=1 comorbidity | 309 (6.2%) | 52 (11.3%) | 361 (6.7%) |
| No/Unknown | 1145 (23.1%) | 117 (25.4%) | 1262 (23.3%) |
| Complications (available data) |  |  |  |
| Yes, >=1 complication | 364 (7.4%) | 119 (25.9%) | 483 (8.9%) |
| No/Unknown | 2363 (47.7%) | 186 (40.4%) | 2549 (47.1%) |

Values are n (%).

### Temporal Trend in Case Fatality

Annual case fatality ranged from 2.17% (2012) to 19.30% (2020) over the study period, with no sustained pattern of increase or decline (Figure 1). The trend analysis showed no significant annual percent change between 2012 and 2025 (slope = +0.254 percentage points/year; p = 0.372). The case fatality peak observed in 2020 coincided with the start of the COVID-19 pandemic; given the small number of cases that year (n = 114), we interpret this figure cautiously in the discussion.

**Figure 1.**
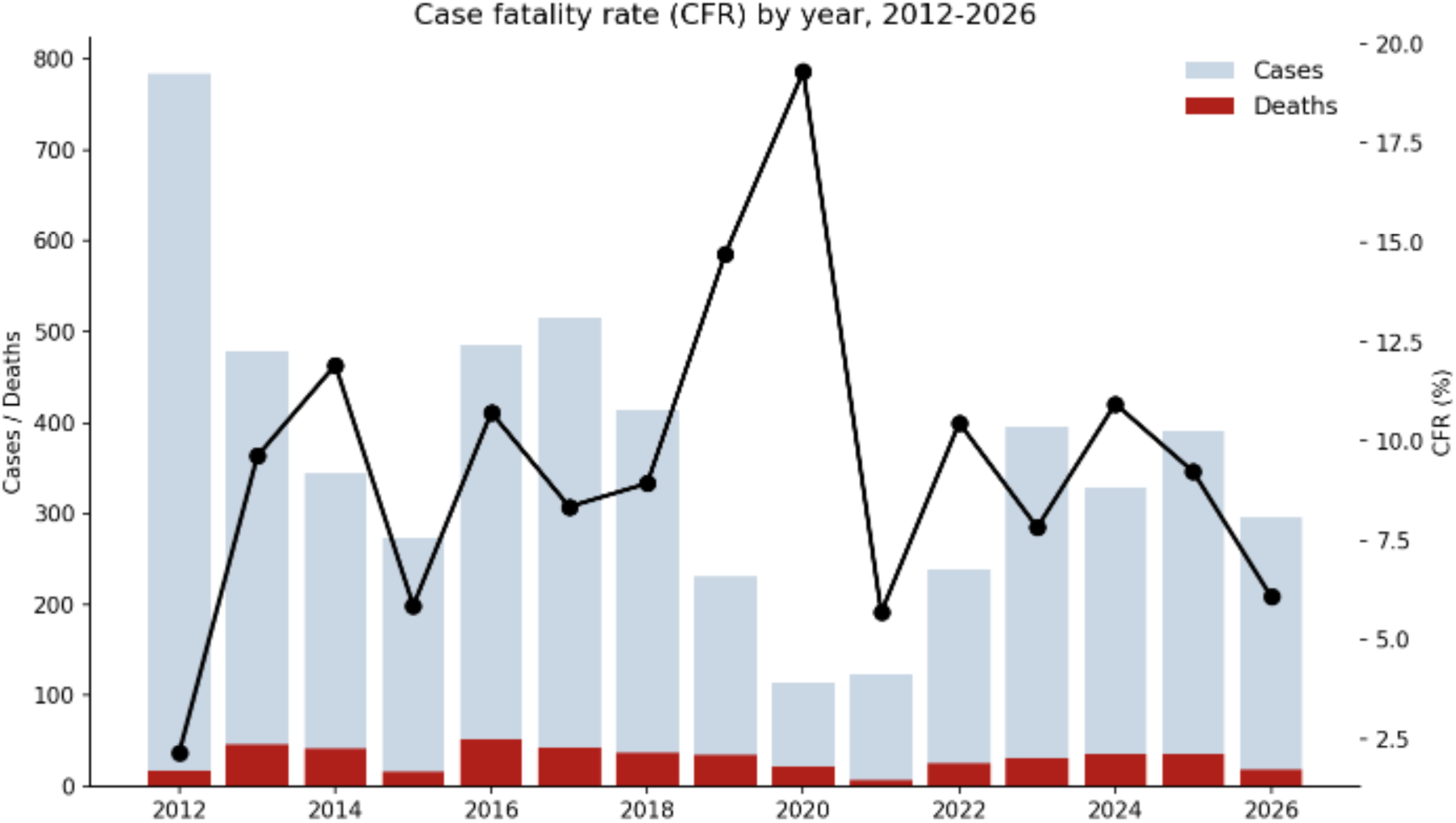
National case fatality by year, with case and death counts.

### Case Fatality by Gender, Classification, and Age

Case fatality was significantly higher in men than in women (9.63%, 95% CI 8.76–10.58 versus 5.49%, 95% CI 4.44–6.78; chi-square, p < 0.001), with a male:female risk ratio of 1.75 (Table 2). Cases classified as probable had the highest case fatality (11.33%), higher than confirmed (8.99%) and suspected cases (7.24%; p < 0.001). We interpret this latter finding cautiously, since it is possible that a proportion of fatal cases die before completing the laboratory confirmation process, remaining classified as probable on clinical-epidemiological grounds rather than confirmed.

**Table 2.** Case fatality by gender, final classification, and age group.

| Dimension | Category | Cases | Deaths | CFR (%) | 95% CI low | 95% CI high |
| --- | --- | --- | --- | --- | --- | --- |
| Gender | Female | 1475 | 81 | 5.49 | 4.44 | 6.77 |
| Gender | Male | 3937 | 379 | 9.63 | 8.74 | 10.59 |
| Classification | Confirmed | 523 | 47 | 8.99 | 6.83 | 11.75 |
| Classification | Probable | 1448 | 164 | 11.33 | 9.79 | 13.06 |
| Classification | Suspected | 3441 | 249 | 7.24 | 6.42 | 8.15 |
| Age | <1 year | 25 | 1 | 4 | 0.71 | 19.54 |
| Age | 1 to 4 years | 169 | 1 | 0.59 | 0.1 | 3.28 |
| Age | 5 to 9 years | 306 | 6 | 1.96 | 0.9 | 4.21 |
| Age | 10 to 19 years | 1299 | 90 | 6.93 | 5.67 | 8.44 |
| Age | 20 to 29 years | 1226 | 132 | 10.77 | 9.15 | 12.63 |
| Age | 30 to 39 years | 718 | 72 | 10.03 | 8.04 | 12.44 |
| Age | 40 to 49 years | 566 | 42 | 7.42 | 5.54 | 9.88 |
| Age | 50 to 59 years | 510 | 57 | 11.18 | 8.73 | 14.21 |
| Age | >60 years | 593 | 59 | 9.95 | 7.79 | 12.62 |

*Chi-square test. Gender: $\chi^2=23.06$ , $p<0.001$ , male:female risk ratio=1.75. Classification: $\chi^2=22.09$ , $p<0.001$ . Age: $\chi^2=52.59$ , $p<0.001$ .*

Case fatality increased significantly with age (chi-square, p < 0.001), from 0.59–4.00% in groups under 10 years to a range of 6.93–11.18% in groups aged 10 and older, without a strictly monotonic pattern: the 20–29 year (10.77%) and 50–59 year (11.18%) groups recorded the highest case fatality, even higher than those over 60 (9.95%) (Table 2).

### Geographic Distribution of Case Fatality

Case fatality varied considerably across the 32 provinces, ranging from 2.43% to 22.22%, and this range narrowed to 5.84–9.90% when aggregated by the eight SNS health regions (Figure 2). Comparing this pattern with the province-level incidence pattern reported for the same period (23), we found a negative correlation between the two measures at the province level (Spearman correlation, r = −0.329; p = 0.066). The highest-incidence provinces did not coincide with the highest-case-fatality provinces, a pattern we discuss as suggestive of differential ascertainment of mild cases across provinces, rather than a true difference in disease severity.

**Figure 2.**
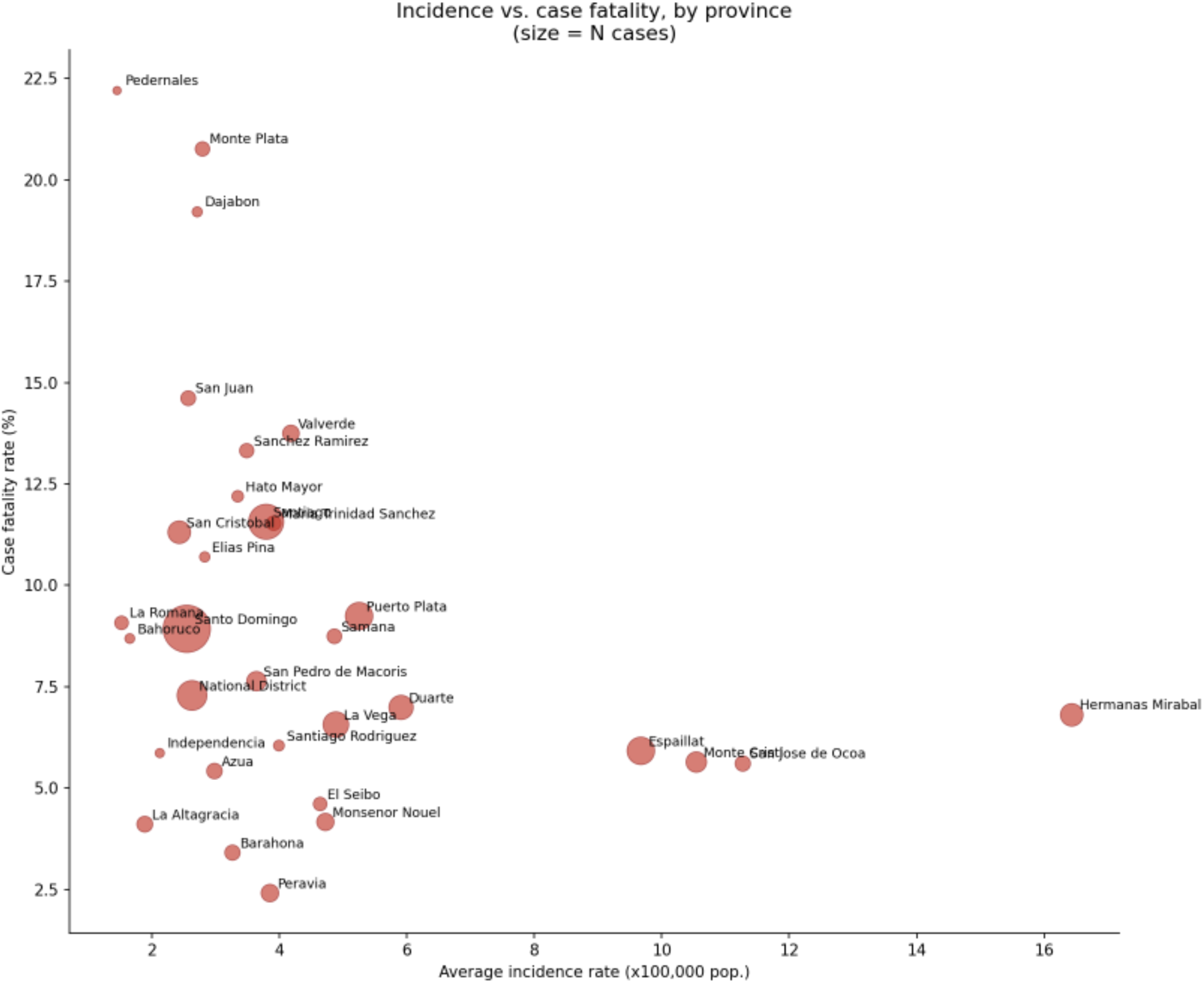
Incidence vs. case fatality by province (geographic triangulation).

### Clinical and Demographic Factors Associated with Case Fatality

In univariate comparisons, case fatality was 24.64% among cases with at least one recorded clinical complication versus 7.30% among those without (chi-square = 133.05, p < 0.0001), and 14.40% among cases with at least one comorbidity versus 9.27% without (chi-square = 7.39, p = 0.0066). In Model A of the multivariable logistic regression (gender, age, comorbidity, and year), male gender (odds ratio [OR] = 1.79; 95% CI 1.18–2.73; p = 0.007) and pre-existing comorbidity (OR = 1.76; 95% CI 1.21–2.57; p = 0.003) were independently and significantly associated with case fatality (Table 3). When clinical complications were incorporated in Model B, they emerged as the strongest predictor of case fatality (OR = 3.16; 95% CI 2.17–4.61; p < 0.0001), while the gender effect attenuated to a marginal level (p = 0.067), suggesting that part of the excess male case fatality may be mediated by a higher frequency of complications in men.

**Table 3.** Multivariable logistic regression: predictors of case fatality (Models A and B).

| Variable | OR | 95% CI low | 95% CI high | p value |
| --- | --- | --- | --- | --- |
| Intercept | 0.09 | 0.05 | 0.15 | <0.0001 |
| Male gender (ref: female) | 1.79 | 1.18 | 2.73 | 0.0067 |
| Comorbidity present (ref: absent) | 1.76 | 1.21 | 2.57 | 0.0034 |
| Year (centered) | 1 | 0.95 | 1.05 | 0.971 |
| Age 0-9 years (ref: 20-29) | 0.06 | 0.01 | 0.44 | 0.0056 |
| Age 10-19 years (ref: 20-29) | 0.58 | 0.35 | 0.97 | 0.0386 |
| Age 30-39 years (ref: 20-29) | 0.89 | 0.54 | 1.47 | 0.6533 |
| Age 40-49 years (ref: 20-29) | 0.72 | 0.41 | 1.27 | 0.2588 |
| Age 50-59 years (ref: 20-29) | 0.88 | 0.51 | 1.51 | 0.6479 |
| Age >60 years (ref: 20-29) | 0.56 | 0.32 | 0.98 | 0.0426 |

***Model B: includes clinical complications (N=1,398)***
| Variable | OR | 95% CI low | 95% CI high | p value |
| --- | --- | --- | --- | --- |
| Intercept | 0.08 | 0.04 | 0.13 | <0.0001 |
| Male gender (ref: female) | 1.53 | 0.97 | 2.4 | 0.0665 |
| Comorbidity present (ref: absent) | 1.58 | 1.05 | 2.39 | 0.0291 |
| Complication present (ref: absent) | 3.17 | 2.18 | 4.61 | <0.0001 |
| Year (centered) | 1 | 0.95 | 1.06 | 0.9361 |
| Age 0-9 years (ref: 20-29) | 0.08 | 0.01 | 0.58 | 0.0126 |
| Age 10-19 years (ref: 20-29) | 0.58 | 0.33 | 1.02 | 0.0597 |
| Age 30-39 years (ref: 20-29) | 0.86 | 0.5 | 1.48 | 0.5901 |
| Age 40-49 years (ref: 20-29) | 0.84 | 0.46 | 1.52 | 0.5661 |
| Age 50-59 years (ref: 20-29) | 0.86 | 0.48 | 1.55 | 0.6169 |
| Age >60 years (ref: 20-29) | 0.56 | 0.3 | 1.02 | 0.056 |

Multivariable logistic regression, Wald test. Model A: male gender $p=0.007$ , comorbidity $p=0.003$ . Model B: complications $p<0.0001$ , strongest predictor.

### Care Timeliness

Time from symptom onset to first medical care did not differ significantly between those who died and survivors (median of 5 days in both groups; Mann-Whitney test, p = 0.078). Fatal cases, however, had slightly faster notification to the surveillance system than survivors (median of 7 versus 8 days; p < 0.0001), a pattern we interpret as reflecting greater clinical urgency in notifying severe cases rather than evidence that delayed notification contributes to fatal outcomes.

## Discussion

Taken together, these results describe an overall case fatality of leptospirosis infection in the Dominican Republic of 8.5% with no significant temporal trend, consistently concentrated among men, in pre-existing comorbidity, and, above all, in the presence of clinical complications. The geographic mismatch between incidence and case fatality, together with the clinical severity field limitation, points to data completeness as a constraint on interpretation, developed further below.

The case fatality rate we detected (8.5%) falls within the 5–15% range classically described for Weil’s syndrome (4–5) and closely matches the case fatality reported in the Colombian analysis of laboratory-confirmed cases for 2015–2020, also 8.5% (22). We consider this convergence particularly informative, given that both countries share a mandatory laboratory surveillance system with a similar structure, suggesting that this figure may reasonably reflect the typical performance of such surveillance systems in the region, rather than a peculiarity of either country. Our figure is considerably lower than the case fatality exceeding 50% described for severe pulmonary hemorrhage syndrome (4–5), consistent with most of our cohort not reaching that extreme of clinical severity.

The finding of significantly higher case fatality in men (risk ratio 1.75) reproduces the demographic pattern consistently described at the global, regional, and comparable-country levels reviewed here: the burden of leptospirosis, in both morbidity and mortality, falls disproportionately on adult men, with global estimates attributing up to 80% of disability-adjusted life years lost to this group (6), a pattern replicated in the Brazilian urban cohort studies (13–18), a recent national Brazilian mortality analysis (24), the Colombian severity analysis (22), and the Thai cohort study (20). This pattern likely reflects greater occupational exposure (agriculture, livestock handling, work in rural or flood-prone areas) and possibly differences in health-care-seeking behavior between sexes, though our data cannot distinguish between these mechanisms.

Pre-existing comorbidity and, above all, documented clinical complications emerged as the strongest predictors of mortality in our multivariable models, consistent with the clinical prognosis literature reviewed here: pulmonary hemorrhage, complicated acute renal failure, and multiple organ failure were the leading causes of death in the Thai cohort (20), consistent with a more recent Thai multicenter study identifying older age, bilirubin, and leptospiremia as independent predictors of in-hospital death (25); a subsequent Thai case-control study confirmed acute renal failure and respiratory failure as leading severity predictors (26); a Malaysian multicenter study identified a similar complication profile among severe cases (27), corroborated by a more recent Malaysian cohort identifying age, delayed hospitalization, and organ-specific markers as independent predictors (28); a Taiwanese cohort similarly identified hemorrhage as an independent mortality predictor (29); and the Colombian analysis found a virtually identical complication profile among severe cases (22), consistent with a recent systematic review of severity predictors (30).

One finding that warrants careful interpretation is the higher case fatality observed among cases classified as probable (11.33%) compared with laboratory-confirmed cases (8.99%). We consider it unlikely that this pattern reflects a true biological difference in disease virulence; we find it more plausible that a proportion of fatal cases die before completing the diagnostic confirmation process, remaining classified on clinical-epidemiological grounds. This interpretation is consistent with the limited confirmatory diagnostic capacity repeatedly documented in the literature reviewed here, both globally (9,21) and specifically in the Caribbean (12,19), and underscores a methodological issue shared with other countries with similarly structured surveillance: final case classification can depend as much on the severity and speed of clinical progression as on the effective availability of confirmatory testing at the appropriate time.

The geographic divergence we documented between the incidence and case fatality patterns by province (the highest-incidence provinces were not the highest-case-fatality provinces, and vice versa) reinforces this same differential-ascertainment hypothesis, and finds a direct parallel in the findings of the Dominican geospatial analysis by Martin and colleagues, which associated higher-risk clusters with distance from health facilities and indicators of structural poverty (11), conditions that plausibly also limit access to confirmatory testing and, therefore, the ascertainment of mild cases. We interpret that provinces with higher recorded incidence likely have better access to diagnostic testing and therefore capture a larger share of mild cases, diluting observed case fatality; whereas in provinces with lower recorded incidence, it is possible that only the most severe cases come to be diagnosed and reported, artificially inflating the case fatality calculated on that basis.

Our study has strengths and limitations that should be considered when interpreting these results. Among its strengths is its national coverage and extended observation period (fourteen complete years plus a partial half-year), which distinguish it from prior studies in the country, focused on two provinces and a single survey year (11,19), and which directly address the geographic and temporal coverage gap flagged by the most recent scoping review of the region (12). Among its limitations, the database does not contain a cause-of-death or date-of-death variable, precluding survival analysis and the distinction between deaths directly attributable to leptospirosis and those due to concurrent causes. The 2020 peak (19.30%) rests on only 114 cases and coincided with COVID-19-related health system strain, and should not be read as a genuine severity spike. Passive surveillance is also subject, as elsewhere in the region and as consistently documented by the underdiagnosis studies reviewed here (9,21), to underreporting and a likely overrepresentation of the most severe cases, which may have inflated observed case fatality relative to the true case fatality among all infected persons.

From a public health perspective, these results suggest concrete lines of action. The comorbidity and clinical complications identified as predictors of mortality offer a clear target for early, aggressive clinical management protocols. The geographic divergence between incidence and case fatality points to specific provinces where strengthening diagnostic capacity, possibly incorporating molecular testing to complement serology as explored in other resource-limited settings (21), could improve early detection of mild cases and potentially reduce observed case fatality.

## Conclusion

In this 14-year national cohort, leptospirosis case fatality in the Dominican Republic was 8.5%, a figure remarkably convergent with that reported in Colombia through a similarly structured surveillance system, with no significant temporal trend, and consistently concentrated among men, among people with pre-existing comorbidity, and, above all, among those who developed clinical complications during the course of illness, a pattern that consistently replicates what is described in the global, regional, and comparable-country literature. The lack of correspondence between the geographic patterns of incidence and case fatality, consistent with the health-service-access determinants already identified in prior geospatial studies in the country, suggests that the diagnostic capacity available in each province substantially shapes both case detection and calculated case fatality. These findings, generated from the first national-coverage, long-term case fatality analysis conducted in the country, provide evidence that can help prioritize clinical and epidemiological surveillance resources against a disease whose burden is expected to grow as extreme weather events intensify across the Caribbean.

## Data Availability

The surveillance data analyzed in this study were provided by the Dominican Republic's Ministry of Public Health (MSP), through the General Directorate of Epidemiology (DIGEPI). Because these are national surveillance data containing potentially sensitive epidemiological detail, they are not publicly deposited; de-identified data may be made available by the corresponding author or by DIGEPI/MSP upon reasonable request and subject to institutional approval.

## Declarations

### Ethics Approval and Consent to Participate

This study was reviewed by the Florida Atlantic University Institutional Review Board (protocol IRB2606182, “Leptospirosis in the Dominican Republic, 2000–2026: National Surveillance Trends”), which determined it exempt from federal regulation under 45 CFR 46.104(2)(ii) on June 22, 2026, as a secondary analysis of de-identified national surveillance data. Individual informed consent was not required, consistent with this exemption determination.

### Consent for Publication

Not applicable: this manuscript does not include identifiable individual data, images, or case details.

### Authors’ Contributions

JJS contributed to data curation, formal analysis, methodology, and writing – original draft. LVA contributed to conceptualization, supervision, and writing – review and editing. DDL contributed to data curation, investigation, and writing – review and editing. OAA contributed to validation and writing – review and editing. MBB contributed to investigation and writing – review and editing. JLCR contributed to data curation, resources, and writing – review and editing. TDVD contributed to conceptualization, supervision, project administration, and writing – review and editing. All authors read and approved the final manuscript.

## Acknowledgments

We thank the General Directorate of Epidemiology (DIGEPI) of the Dominican Republic’s Ministry of Public Health for providing access to the national surveillance data that made this study possible.

## Declaration of Generative AI and AI-Assisted Technologies

During the preparation of this manuscript, the authors used Claude to assist with language editing, clarity, and readability. Claude was not used for data analysis, statistical modeling, interpretation of results, or the generation of scientific conclusions. All AI-assisted content was reviewed and revised by the authors, who take full responsibility for the accuracy, integrity, and final content of the manuscript.

